# Endometriosis Online Communities: A Quantitative Analysis

**DOI:** 10.1101/2024.02.27.24303445

**Authors:** Federica Bologna, Rosamond Thalken, Kristen Pepin, Matthew Wilkens

**Affiliations:** Department of Information Science, Cornell University; Department of Obstetrics and Gynecology, Weill Cornell Medical College

**Keywords:** online health communities, patient-centered care, chronic disease, internet, consumer health information, self-help groups, community networks, information science, social support

## Abstract

**Background:** Endometriosis is a chronic condition that affects 10% of people with a uterus. Due to the complex social and psychological impacts caused by the condition, people with endometriosis often turn to online health communities (OHCs) for support.

**Objective:** Prior work identifies a lack of large-scale analyses of endometriosis patient experiences and of OHCs. Our study fills this gap by investigating aspects of the condition and aggregate user needs that emerge from two endometriosis OHCs, *r/Endo* and *r/endometriosis*.

**Methods:** We leverage topic modeling and supervised machine learning to identify associations between a post’s subject matter (“topics”), the people and relationships (“personas”) mentioned, and the type of support the post seeks (“intent”).

**Results:** The most discussed topics in posts are *medical stories, medical appointments, sharing symptoms, menstruation*, and *empathy*. In addition, when discussing *medical appointments*, users are more likely to mention the *endometriosis OHCs* than *medical professionals*. Furthermore, *medical professional* is the least likely of any persona to be associated with *empathy*. Posts that mention *partner* or *family* are likely to discuss topics from the *life issues* category, in particular *fertility*. Lastly, we find that while users seek experiential knowledge regarding treatments and healthcare processes, they also wish to vent and to establish emotional connections about the life-altering aspects of the condition.

**Conclusions:** Endometriosis OHCs provide members a space where they can discuss care pathways, learn to manage symptoms, and receive validation. Our results emphasize the need for greater empathy within clinical settings, easier access to appointments, more information on care pathways, and further support for patient loved ones. In addition, this study demonstrates the value of quantitative analyses of OHCs: they can support and extend findings from small-scale studies about patient experiences and provide insight into hard-to-reach groups. Lastly, analyses of OHCs can help design interventions to improve care, as argued in previous studies.

## Introduction

### Endometriosis

Endometriosis is a chronic condition that affects 10% of people with a uterus and is characterized by the presence of uterine lining tissue outside of the uterus [1]. This condition causes a range of painful, persistent, and life-altering symptoms, including, but not limited to chronic pelvic pain, painful menstruation, constipation, painful urination, painful sexual intercourse, and infertility. There is no cure for endometriosis, so treatment focuses on symptom management and relief [1–3]. Treatments might include hormonal therapy, surgical removal of endometriosis, and fertility treatment. However, such therapies cause numerous side effects and rarely provide long-term relief to patients [2].

Due to the absence of condition-specific symptoms and biomarkers, the normalization of menstrual pain, the need for surgery to make a diagnosis, and the lack of knowledge about the condition by both the public and clinicians, the average time until diagnosis is estimated to be between 6 to 11 years, depending on the healthcare system of reference [4–6]. A confirmed diagnosis can only be reached through laparoscopic excision of endometriosis, an invasive surgical procedure [7].

Endometriosis patients face numerous difficulties during their healthcare journeys. Not only do they struggle to find information, but they also encounter negative attitudes from physicians [8–10]. Patients’ concerns are often dismissed as ‘just period pain’ by providers [11]. Negative attitudes seem to derive from physicians’ own discomfort with unexplained symptoms [12], as well as from the continued presence of hysteria discourse and androcentric views in medical literature [13].

Because of these interconnected factors, endometriosis has dire impacts on patients’ quality of life [14]. The condition forces people to leave their education and employment and to opt out of social events and everyday activities. Due to sexual pain and infertility, patients may feel inadequate as partners and fear abandonment [15].

Endometriosis patients necessitate support from partners, family members, and friends to overcome these struggles and to receive a diagnosis [6,16]. Self-care practices are time-consuming and labor-intensive for both the patients and their loved ones. As patients focus on following complex treatment regimens and become experts in their own care [17–20], a wide range of responsibilities falls onto partners and family members. These responsibilities can include financial and housekeeping duties, helping to navigate the healthcare system, and relaying medical information, among others [21–24].

Research on endometriosis highlights several areas of endometriosis care that require improvement. Medical treatments should be more holistic, taking into consideration the social, emotional, and psychological costs of endometriosis for sufferers and their loved ones [1,15,16]. Health care providers should improve their communication to validate patients’ concerns, meet their informational needs, and avoid misunderstandings [5,9,18]. Since loved ones are also affected by the condition, they should receive education and training on the condition from healthcare professionals [15,25–27].

### Online Health Communities

OHCs are groups of individuals who come together on an Internet-based platform (e.g., social media, website, or forum) to discuss general or condition-specific medical topics. Members may be patients, medical professionals, informal caregivers, patients’ loved ones, or members of the general public [28,29].

OHCs have been shown to provide support to users who experience dissatisfaction or constrained access to medical care, limited social support, or the absence of a local community of people with the same condition [21]. Indeed, some members join OHCs after feeling alienated from the medical community, or becoming distrustful of medical knowledge and care [28,29]. Others join to learn about alternative treatment options, or to advocate for better awareness of their condition [30,31].

As these communities allow for varying levels of pseudonymity and anonymity, users with stigmatic and chronic conditions can share intimate or stigmatized information without fearing social repercussions [32,33]. People with chronic conditions often use OHCs to make sense of their experiences and receive validation [34,35].

Studies of OHCs show that an individual member’s support needs may change over time [36,37]. Earlier work on support matching suggests that different types of support may be more appropriate for certain needs [38]. A study of a breast cancer OHC found that the presence of emotional or informational support increased the original poster’s satisfaction, though users expressed less satisfaction if they received emotional support when seeking informational support [39]. A separate study on a mental health OHC found that support matching positively predicted satisfaction, but that there was significant variance across users [36].

Participating in OHCs empowers members as they become better informed about their health concerns, learn to manage their condition, and gain strategies for communicating with healthcare providers [10,28,40–44]. In many cases, they ultimately feel less isolated. Contrary to common belief, Huh finds that OHC members do not share misinformation and commonly invite peers to consult a provider for medical advice [45]. Other studies of OHCs confirm the beneficial effects of engaging in these communities, showing that members gradually express more positive emotions than negative ones with sustained participation [46,47].

One study of an addiction recovery OHC found that engagement in the community correlates positively with recovery [48].

Researchers have also highlighted OHCs’ role in the improvement of healthcare [49,50]. An existing study of a PCOS subreddit found concordance between trends from lab results posted to the OHC and trends from clinical research. This indicates that, although OHCs often include patients that are typically excluded from clinical trials (such as those with multiple conditions), studying these communities is useful to understand patient populations [51]. Indeed, content analysis of these communities reveals patterns across patients’ experiences of care and symptoms [52–55] and OHC members’ expertise in providing support to peers could be leveraged to deliver healthcare interventions and programs [41,43,50,56].

### Endometriosis Online Communities

Due to the significant impacts of the condition on patients’ lives, people suffering from endometriosis often turn to both offline and online communities for help. The former generally consist of dedicated in-person meetings and activities, and access depends on proximity [57,58]. The latter exist in a variety of forms, such as blogs, mailing lists, Facebook pages, and Instagram accounts; their activities depend on the specific platform [28,34].

Whelan et al. find that both an offline and an online endometriosis group are epistemic communities. As members share their stories and interact with peers, they build a new epistemology in which patient experiences become valid forms of knowledge [59].

Previous research also focuses on the kinds of support and content shared in endometriosis online communities. In a study of Facebook pages for people with endometriosis, Towne et al. show that 48% of posts provided emotional support, while educational posts made up 21% of the total. Furthermore, they find that 94% of the educational posts shared accurate information [60]. On the other hand, Metzler et al. find that most posts on Facebook and Instagram accounts about endometriosis offer inspiration or support, awareness about the disease, or personal information. Followers mostly engage with posts that are humorous, generate awareness, and contain personal content [61]. Finally, Shoebotham and Coulson demonstrate that several therapeutic benefits are related to joining endometriosis online support groups. They find that members feel reassured and empowered while improving their knowledge of endometriosis [62].

### Contribution

Prior work identifies a lack of large-scale mixed-method analyses on endometriosis patient experiences and on online communities. It specifically calls for studies regarding:

- what is discussed in endometriosis online communities [61];
- the impact of endometriosis on loved ones and informal caregivers [15];
- the impact of endometriosis on adolescents [15];
- the impact of endometriosis-induced infertility [15];

Indeed, existing qualitative research on patient experiences has been limited to small patient samples. In contrast, quantitative analyses have used ontologies defined by researchers, rather than inferred from patient narratives [15,60,61]. Furthermore, most studies on the effects of endometriosis on quality of life only include people with an endometriosis diagnosis within the research population (e.g., [8,9,11,18,63]). Given the long average delay between symptom onset and diagnosis [1,5], many people with endometriosis are missed by this research.

This study fills this gap by providing a large-scale analysis of user behavior in two endometriosis OHCs, *r/Endo* and *r/endometriosis*. By studying these communities, we can discover the unmet needs of hard-to-reach groups. Thanks to the pseudonymity afforded by the platforms, users feel more comfortable discussing needs that they might not have the time or courage to address in clinical settings. In addition, these OHCs are open and accessible to anyone, regardless of whether they have a diagnosis or not. As a result, numerous members belong to populations that have been missed by endometriosis research: people who are pre-diagnosis, adolescents, and loved ones of people with a diagnosis.

Using natural language processing, we identify and map the associations between a post’s subject matter (“topics”), the people and relationships (“personas”) mentioned, and the type of support the post seeks (“intent”). We investigate two research questions:

- RQ1: What aspects of the endometriosis experience are discussed in OHCs?
- RQ2: What aggregate needs emerge from the OHCs?

## Methods

### Data

Endometriosis OHCs exist on many platforms in many forms [28,34]. We study two thriving endometriosis subreddits, *r/Endo* and *r/endometriosis*, which feature high membership and participation numbers (Table 1), and show promise of continued growth (Figure 1-2). We collect posts and comments from *r/Endo* and *r/endometriosis* from their inception (January 2012 and November 2014, respectively) to December 2021 using the Pushshift Reddit API. We make available the custom Python code used for the data collection process and subsequent analysis.

**Table 1.** General statistics of r/Endo, r/endometriosis, and of the combined dataset.

|  | *r/Endo* | *r/endometriosis* | combined |
| --- | --- | --- | --- |
| Number of posts | 22,584 | 12,131 | 34,715 |
| Number of comments | 225,221 | 127,941 | 353,162 |
| Number of members | 40,734 | 38,270 | 79,004 |
| Unique posters | 20,262 | 17,150 | 20,263 |
| Mean number of words per post | 184 | 182 | 184 |
| Mean number of words per comment | 67 | 66 | 67 |
| Mean number of comments per post | 8 | 9 | 9 |
| Total number of words | 19,363,897 | 10,606,139 | 29,970,036 |
| Number of unique tokens | 110,055 | 76,379 | 138,106 |

**Figure 1.**
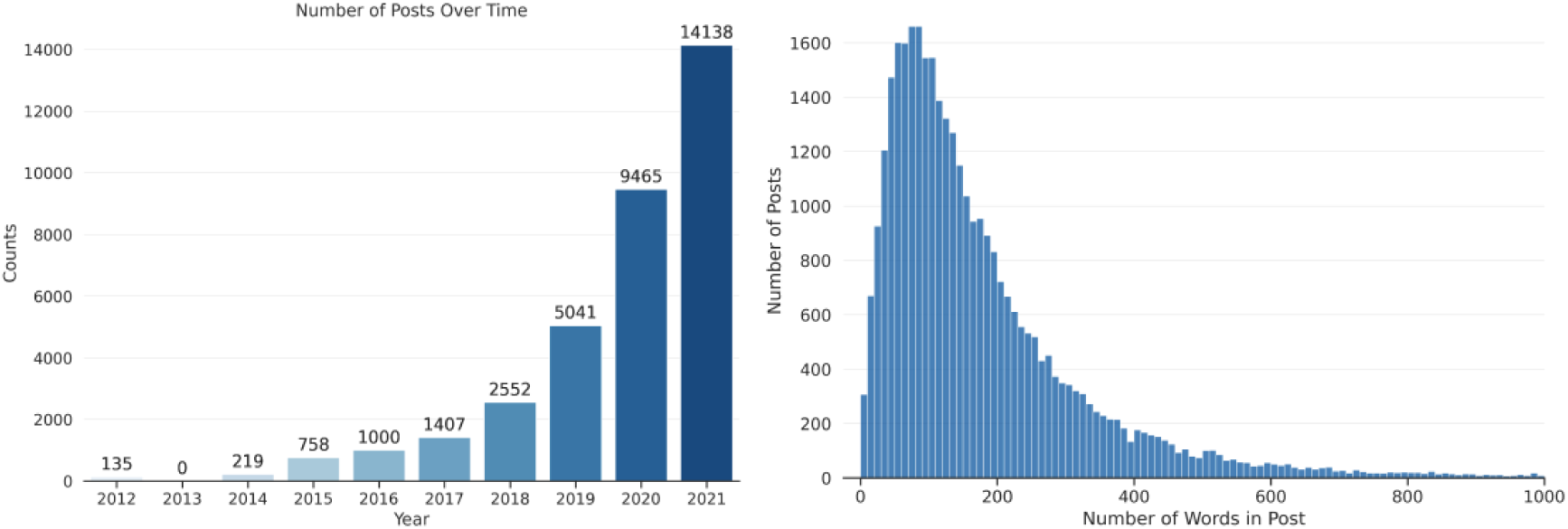
Number of posts in r/Endo and r/endometriosis over time (left) and distribution of post lengths by number of words (right).

**Figure 2.**
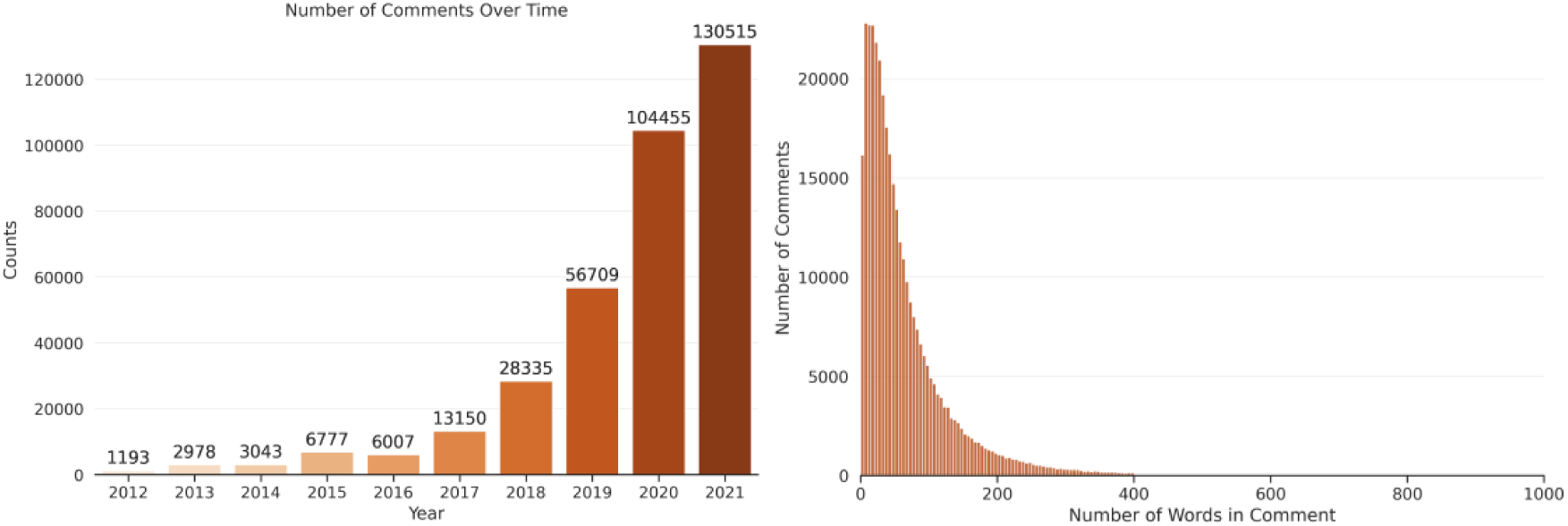
Number of comments in r/Endo and r/endometriosis over time (left) and distribution of comments’ lengths by number of words (right).

After reading posts, examining general statistics of the subreddits (Table 1), and comparing their community-specific languages using Monroe et al.’s Fightin’ Words method [64] (available in the Appendix A), we find that the two communities share sufficient similarities to justify treating them as a single dataset.

#### Ethical Framing

People with endometriosis have historically been failed by research and medical institutions. Like other gendered conditions, compared to its severity and the number of people diagnosed with it, endometriosis is greatly underfunded [65]. There is a persistent imbalance between the high percentage of people with endometriosis and the low number of endometriosis experts [1]. Patients deal with ongoing disbelief, invalidation, or trivialization of their symptoms, even from members of the medical community [8,10,13]. As academic researchers who are not members of the endometriosis community, it is imperative that we handle users’ data with care.

Though data from *r/Endo* and *r/endometriosis* is public, members of online communities do not necessarily anticipate that their posts and comments could be used by academic researchers [66]. By collecting, analyzing, and publishing research about this data, we extract the data from its intended audience, bringing it to a new, unanticipated audience [67]. Following prior examples of handling sensitive, health-related data [30,52,68], we obscure the source data to protect members from being identified in relation to their posts or comments. Obfuscation is performed in two ways: 1) throughout this work, we paraphrase any quoted material and 2) we do not re-release the underlying text data itself. Any quoted material in the paper has undergone rewording at the sentence level to make it less directly searchable, but we retain as much content of the original version as possible. We release all code and our codebooks so that other researchers may replicate our results on future versions of the OHC, subject to users’ later in situ modifications or deletions of their contributions.

### Computational Text Analysis

We use complementary supervised and unsupervised methods to isolate specific instances of personas and intents, but also to allow topics to emerge beyond the research questions we have designed.

#### Topic Modeling

Following suggestions from research on endometriosis experiences [15], we extract topics from the two endometriosis OHCs using an abductive approach, rather than a priori categories. First, we extract topics from posts and comments using unsupervised topic modeling. Successively, we evaluate our list of topics against themes previously identified in qualitative research.

To extract topics from our collection of posts and comments, we use latent Dirichlet allocation (LDA) [69], a type of statistical topic modeling. For each topic in the model, every individual word in the collection is assigned a probability of belonging to a given topic. Consequently, each document (e.g., a post or comment) is assigned a higher or lower probability of representing each topic depending on the words it features.

Before training the LDA model, we clean posts and comments using the string processor included in Antoniak’s `little-mallet-wrapper` [70], which is designed to prepare raw text for topic modeling. The string processor splits strings into a series of tokens (words separated by punctuation or spaces), removes punctuation and common words, converts all characters to lowercase, and returns the transformed string. After this initial cleaning, we remove any post and comment written by or responding to bots by searching for the string ‘bot’ in both the user name and the text of the document. Next, we implement the LDA function using the `tomotopy` Python package [71].

We experiment by running multiple models with different combinations of the following parameters: number of topics=10,15,20,25; number of removed most frequent words=5,10,15,20. We also explore training the model with different document lengths. We first run LDA on whole posts and comments, then we chunk these into paragraphs and sentences.

To evaluate the performance of each model, we read each topic’s top 100 documents by average probability and assign a descriptive label to each topic based on the content of those documents.

Following this evaluation procedure, we find the topic model trained with 25 topics on paragraph chunks to best suit our purposes. We group topics into 5 overarching categories based on conceptual similarity and interconnectedness: *symptoms, medications, healthcare, self-care,* and *life issues*. The detailed description and listing of the 5 categories, the 25 topics, and each topic’s top 10 keywords is shown in section 5.1.1 below. We then compare our topics against themes identified in previous research on endometriosis.

#### Supervised Classification

As a complement to the unsupervised topics, we design two supervised tasks: the identification of people based on their social roles (*personas*) in posts and the identification of the goal (*intent*) of a post. Supervised machine learning allows us to assign OHC-specific labels, including personas and intent, to all posts in our dataset.

##### Personas

Personas are types of people, organized by social roles, who often interact with a person with endometriosis. We identify discussions of personas in endometriosis OHC posts to better understand how endometriosis interfaces with interpersonal relationships. Specifically, we study the four most frequent personas mentioned in the endometriosis OHCs, based on a qualitative analysis of 200 posts: *medical professional*, *partners*, *family*, and the *endometriosis OHCs* themselves. Given the variety of terms that could represent each persona (e.g., a gynecologist, a subcategory of *medical professional*, could also be referred to as gyno, obgyn, gynecologist, obstetrician, doctor, doc, provider, or many others), instead of using a keyword search for each persona category, we train a supervised model to identify personas based on hand-labeled examples.

*Medical professional* is any type of professional in the healthcare system with a patient-facing role, such as a doctor, gynecologist, nurse, etc. The *partner* persona includes romantic partners, and *family* includes mentions of family members (e.g., parents, children, siblings). Depending on paragraph context, *family* may also encompass *partners*. The *endometriosis OHCs* label involves the *r/Endo* and *r/endometriosis* subreddit communities. Paragraphs that mention the subreddit might do so by name, but they also include posts that speak directly to the reader (e.g. “can you tell me if you’ve experienced this?”). The *endometriosis OHCs* label differs from the others, given that the endometriosis OHCs tends to be both the audience and subject matter of a post.

In a random sample of paragraphs from posts in the corpus, we assign the paragraph a label for every present persona category. If there is no persona present, the paragraph does not receive a label. To assess inter-rater reliability, using the labeling scheme described above (alongside a codebook included in Appendix B), two authors labeled 200 of the same randomly sampled paragraphs. Using Cohen’s kappa, we reach satisfactory inter-rater reliability across all categories. Then, for each persona, one author labeled paragraphs until reaching enough labeled data for acceptable classification performance, resulting in a different number of total paragraphs labeled for each category (Table 2).

**Table 2.** Number of paragraphs assigned the persona labels out of total paragraphs labeled and inter-rater reliability.

| Persona | Paragraphs Assigned Label / Total Labeled | Inter-Rater Reliability (200 post subset) |
| --- | --- | --- |
| Family | 153/1500 | 0.79 |
| Partner | 166/2000 | 0.83 |
| Medical Professional | 349/1000 | 0.87 |
| Endometriosis OHCs | 368/1000 | 0.84 |

##### Persona Models Setup and Prediction

For each persona category, we fine-tune a pre-trained DistilBERT model on the persona-annotated paragraphs to perform a binary classification task [72]. DistilBERT is an English-language large language model that can be fine-tuned on a given dataset to perform a specific task, such as supervised classification [73]. DistilBERT provides a lightweight version of BERT that retains much of its performance, making it easier for other work to replicate our results and to use our trained models. For each persona category, we fine-tune DistilBERT on paragraphs from both endometriosis OHCs, to best predict the assigned categorical label. We keep all training hyperparameters consistent across models, using a learning rate of 5e-5, 50 warm-up steps, and a weight decay of 0.01, in three training epochs. As a baseline model, we also perform logistic regression on each persona category, with input texts in term frequency - inverse document frequency (TF-IDF) structure. Classification accuracy for a held-out test set of 25% of the total labeled paragraphs is listed in Table 3. For all classification results, we present macro scores, which are a more pessimistic scoring method that treats both classes equally, regardless of class imbalance. We use each trained model to predict instances of personas in paragraphs in the rest of the corpus.

**Table 3.** Classification performance for each persona category, for both logistic regression and DistilBERT. All scores are reported as macro averages.

| Persona | Classifier | Precision | Recall | F1 |
| --- | --- | --- | --- | --- |
| **Family** |  |  |  |  |
|  | Logistic Regression | 0.50 | 0.45 | 0.48 |
|  | DistilBERT | 0.94 | 0.92 | 0.93 |
| **Partner** |  |  |  |  |
|  | Logistic Regression | 0.50 | 0.46 | 0.48 |
|  | DistilBERT | 0.91 | 0.97 | 0.93 |
| **Medical Professional** |  |  |  |  |
|  | Logistic Regression | 0.71 | 0.83 | 0.72 |
|  | DistilBERT | 0.93 | 0.93 | 0.93 |
| **Endometriosis OHCs** |  |  |  |  |
|  | Logistic Regression | 0.72 | 0.83 | 0.73 |
|  | DistilBERT | 0.93 | 0.92 | 0.92 |

##### Intent

Prior research on support in OHCs has established multiple overarching categories of support, often characterized as either emotional or informational support [36,39,74]. OHC research takes these support categories and maps them onto behavioral features in the data, which suggest the type of support a person seeks or provides [37]. Our work specifically considers what users desire from the act of posting, which we call their intent, but we acknowledge that the intent of a post is unavailable to researchers without directly speaking to the person who shared a post. To develop a set of intent categories that are tailored to the endometriosis OHCs, we iteratively label, discuss, and revise our labels. We identify four common categories of intent: *seeking informational support*, *seeking experiences*, *seeking emotional support*, and *venting*.

##### Seeking Informational Support

Seeking informational support occurs when a person posts to the OHC to find medical information. We build upon prior definitions of seeking informational support [37,75], but incorporate a novel but simple heuristic for labeling: could the post’s question be usefully posed to a doctor? After revising the *seeking informational support* definition, we found major improvements in labeling consistency, speed, and inter-rater reliability. Adding this question also created an effective distinction between *seeking informational support* and *seeking experiences*.

> *My gyno said there’s a chance I have endo, but that I can’t be diagnosed yet since I’m too young (21). Is that true? Is there some sort of test I should be pushing for? I had a doctor who refused to perform a pelvic exam because she said I couldn’t have digestive problems because of endo. I’m feeling skeptical and I don’t know how to advocate for myself.*

##### Seeking Experiences

Seeking experiences is the inverse of seeking informational support}, as posts that seek experiences could only be answered by someone exposed to the endometriosis experience or who has been on the receiving end of care. Posts that seek experiences ask the community for their experiences with a variety of medical procedures or their day-to-day experiences living with endometriosis. Some of these posts may also ask if members of the community have experienced similar symptoms.

> *Does this sound like endo? How did you get your diagnosis? Did you go to a specialist? Any other advice is appreciated.*

##### Seeking Emotional Support

Seeking emotional support includes posts that ask for encouragement, empathy, validation, or help navigating emotional situations. These posts may look for emotional support after a negative experience, but they may just as easily ask for celebration from the community after a major milestone in care, such as a diagnosis, improvements in symptoms, or successful self-advocacy.

> *I’m feeling really down and I can’t talk to my doctor. The only reason she agreed to do this was because of my mental illness. I’m so afraid that either outcome will break my heart. How do I live with the results?*

##### Venting

Our final label, venting, occurs when a person posts about their grievances living with endometriosis or frustration at a specific situation. We are not aware of similar labels in previous OHC research. Both communities support the practice of venting or ranting, and even have “flares” (tags) for posts that vent or rant.

> *This is a long post, but I’m feeling hopeless. I started dealing with things since around 12 years old and now I’m 26. This pain has lasted for weeks and I can’t do any of the physical activities that I love and I feel useless and everyone is dismissing me like a crazy person. I feel dismissed by today’s doctor, some woman on the phone, all the doctors I’ve ever dealt with since 12. Ugh sorry I know this is long but I needed to rant. Anyway thanks for listening to me talk it out.*

We find that most posts begin or end by stating the person’s intent and their preferred form of support. Whenever possible, we choose the intent that aligns with a post’s explicitly stated purpose.

Using this codebook (included in Appendix C), one author labeled 1500 sampled posts from *r/Endo* and *r/endometriosis*, to be used as training data for our models. Each post can receive between zero to all four intent category labels, though most posts have a primary, explicitly expressed intent. A second author labeled 200 of the same posts as those used for training the models, to be used for measuring interrater reliability. Using Cohen’s kappa, we reach acceptable inter-rater reliability across all categories (Table 4).

**Table 4.** Number of posts assigned the intent labels out of 1500 posts and inter-rater reliability for each label.

| Label | Posts Assigned Label | Inter-Rater Reliability<br>(200 post subset) |
| --- | --- | --- |
| Seeking Informational Support | 524 | 0.79 |
| Seeking Experiences | 691 | 0.83 |
| Seeking Emotional Support | 241 | 0.76 |
| Venting | 172 | 0.74 |

##### Intent Models Setup and Prediction

We fine-tune a series of DistilBERT models to perform binary classification to predict each intent category in a post. Classification accuracy for a held-out test set of 25% of the total labeled paragraphs is listed in table 5. Overall, the intent models reach acceptable performance, though it is lower than that of our persona models. This slightly lower performance is expected because of the more complex nature of the intent categories. We then use the fine-tuned models to predict the intent of posts in the entire corpus.

**Table 5.** Classification performance for each intent category, for both logistic regression and DistilBERT.

| Intent | Classifier | Precision | Recall | F1 |
| --- | --- | --- | --- | --- |
| **Seeking Informational Support** |  |  |  |  |
|  | Logistic Regression | 0.59 | 0.71 | 0.56 |
|  | DistilBERT | 0.86 | 0.82 | 0.84 |
| **Seeking Experiences** |  |  |  |  |
|  | Logistic Regression | 0.75 | 0.75 | 0.75 |
|  | DistilBERT | 0.83 | 0.83 | 0.83 |
| **Seeking Emotional Support** |  |  |  |  |
|  | Logistic Regression | 0.51 | 0.80 | 0.47 |
|  | DistilBERT | 0.72 | 0.69 | 0.70 |
| **Venting** |  |  |  |  |
|  | Logistic Regression | 0.50 | 0.44 | 0.47 |
|  | DistilBERT | 0.83 | 0.80 | 0.81 |

## Results

### RQ1: What aspects of the endometriosis experience are discussed in OHCs?

Leveraging topic probabilities, we investigate which aspects of endometriosis experiences are discussed in the endometriosis OHCs. We first consider what topics emerge from the endometriosis OHCs once we perform LDA topic modeling on paragraph chunks from posts and comments. Secondly, we analyze which of those topics are the most discussed in posts.

#### Topics in Posts and Comments

Employing LDA topic modeling, we find discussions of five main topic categories in the endometriosis OHCs: symptoms, medications, healthcare, self-care practices and life issues. A complete list of the five categories and our 25 topics is provided below. Although healthcare is the category with the highest number of topics, symptoms and life issues are also largely discussed in these communities. We also find the importance of self-care practices, as they are discussed substantially enough that we place them in a separate category.

##### Symptoms

A major pattern in the two OHCs is the presence of topics related to symptoms (Table 6). People with endometriosis suffer from a wide range of disabling chronic symptoms: *gastrointestinal* issues; *pelvic floor* pain; heavy, irregular, and painful *menstruation*; *muscular* cramps in their legs and abdomen. Many users share these symptoms with the communities in hope of receiving or providing support.

**Table 6.** Topics in the *symptoms* category. Numbers are assigned randomly by the model, while labels are assigned upon reading 100 documents for each topic.

| Topic # | Label | Top 10 words |
| --- | --- | --- |
| 0 | Gastrointestinal | take nausea bowel stomach help water constipation taking drink helps |
| 3 | Pelvic floor | pelvic floor therapy physical help sex helped therapist muscles lot |
| 5 | Menstruation | period periods days bleeding symptoms painful heavy cramps started normal |
| 17 | Muscular | back sex right feel feels lower side sometimes left painful |
| 21 | Sharing symptoms | feel period day days bad time every worse back last |

##### Medications

Due to the chronic nature and current incurability of endometriosis, people with endometriosis make use of a variety of drugs and treatments. Users of the two OHCs often list their *pain management* routine, share *hormonal treatment experiences* (“*18 months ago I started using the Nuva ring and I love it.*”), recount the side effects of specific drugs they have used, or provide medical *information on hormonal drugs* (“*Orlissa is a GnRH antagonist, so it lowers estrogen directly without relying on the same feedback mechanism as Lupron*”). The *medications* category groups these experiences (Table 7).

**Table 7.** Topics in the *medications* category. Numbers are assigned randomly by the model, while labels are assigned by reading the top 100 documents for each topic.

| Topic # | Label | Top 10 words |
| --- | --- | --- |
| 14 | Pain management | work take cbd time day job days help use much |
| 18 | Hormonal drug experiences | control months birth iud pill period years mirena periods got |
| 23 | Drugs | take side taking effects weight months pill medication dose NUMmg |
| 24 | Information on hormonal drugs | control birth symptoms treatment side estrogen effects hormones lupron hormonal |

##### Healthcare

In the *healthcare* category, we group topics regarding the medical aspects of endometriosis, and how endometriosis patients experience the healthcare system (Table 8). Often, senior members of the OHCs provide new users with *medical information* on the condition, overviews on the process of *getting diagnosed*, as well as *information on surgery*. Users also advise each other on how to prepare for their *medical appointments*. They often point to competent endometriosis *specialists*, compare *insurance* policies, and highlight helpful *online resources*.

**Table 8.**
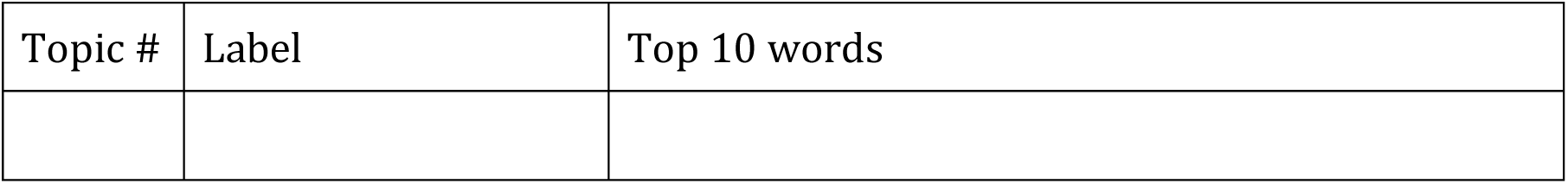
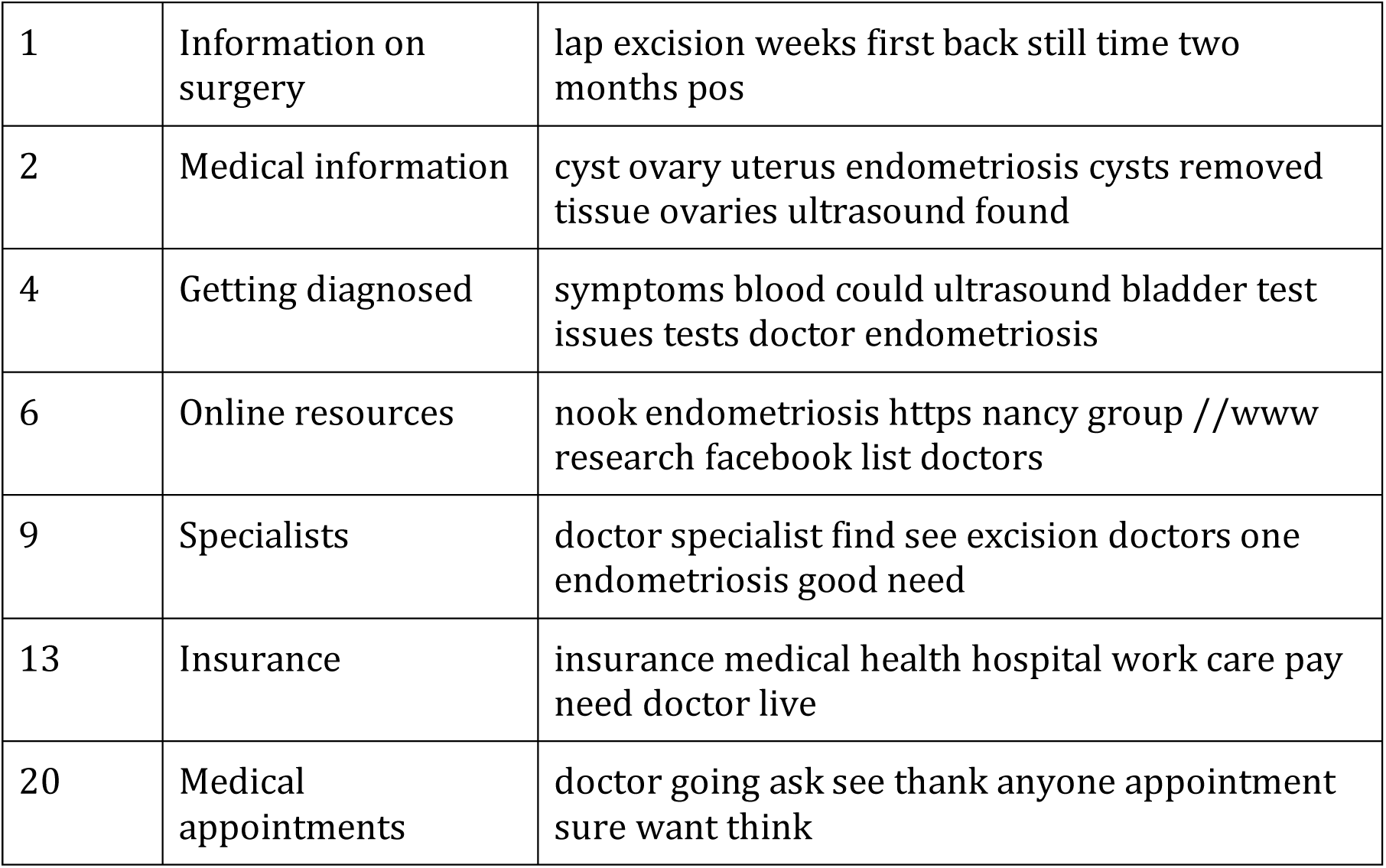
Topics in the *healthcare* category. Numbers are assigned randomly by the model, while labels are assigned upon reading the top 100 documents for each topic.

##### Self-care

As endometriosis requires a considerable amount of *self-care* (Table 9), patients are faced with the challenge of caring for themselves while also having work and other responsibilities. Users of the OHCs find support against exhaustion and isolation by comparing experiences and tips about their *post surgery recovery*. They also provide detailed information on their *diet*, *product recommendations* for gadgets that help with daily activities, and various *comfort items* for when symptoms flare-up.

**Table 9.** Topics in the *self-care* category. Numbers are assigned randomly by the model, while labels are assigned upon reading the top 100 documents for each topic.

| Topic # | Label | Top 10 words |
| --- | --- | --- |
| 8 | Post surgery recovery | day days first home time around back gas hours week |
| 15 | Product recommendations | heating pad use hot heat one water help helps pads |
| 19 | Diet | diet eat gluten food foods dairy eating try free lot |
| 22 | Comfort items | wear pants belly look weight one size wearing super cup |

##### Life issues

The last category, *life issues*, groups users’ discussions of general life issues connected with having a severe chronic condition (Table 10). In these communities, users open up about their experiences of *dismissal and abuse* and their *medical stories* as patients. They give each other support through their *fertility* struggles. Community members exchange expressions of *gratitude* and *empathy* with their peers.

**Table 10.** Topics in the *life issues* category. Numbers are assigned randomly by the model, while labels are assigned upon reading the top 100 documents for each topic.

| Topic # | Label | Top 10 words |
| --- | --- | --- |
| 7 | Dismissal | people even doctors think feel something say one want women |
| 10 | Gratitude | hope thank good much sorry better feel luck well find |
| 11 | Medical stories | years told doctor said got went diagnosed back lap finally |
| 12 | Fertility | pregnant want kids years hysterectomy fertility pregnancy |
| 16 | Empathy | feel people life want help much need support sorry hard |

#### Most Discussed Topics in Posts

To investigate which aspects of endometriosis patient experiences are most discussed in the endometriosis OHCs, we measure which topics have the highest average probability in all posts. Indeed, if a topic shows a high average probability across all posts, it indicates that the topic is highly present in the endometriosis OHCs. In posts, the topics with the highest average probability are *medical stories*, *medical appointments*, *sharing symptoms*, *menstruation* and *empathy* (Table 11, Figure 3).

**Table 11.** Topics with highest average probability in posts.

| Topic # | Label | Average probability |
| --- | --- | --- |
| 11 | Medical stories | 0.086 |
| 20 | Medical appointments | 0.081 |
| 21 | Sharing symptoms | 0.080 |
| 5 | Menstruation | 0.079 |
| 16 | Empathy | 0.067 |

**Figure 3.**
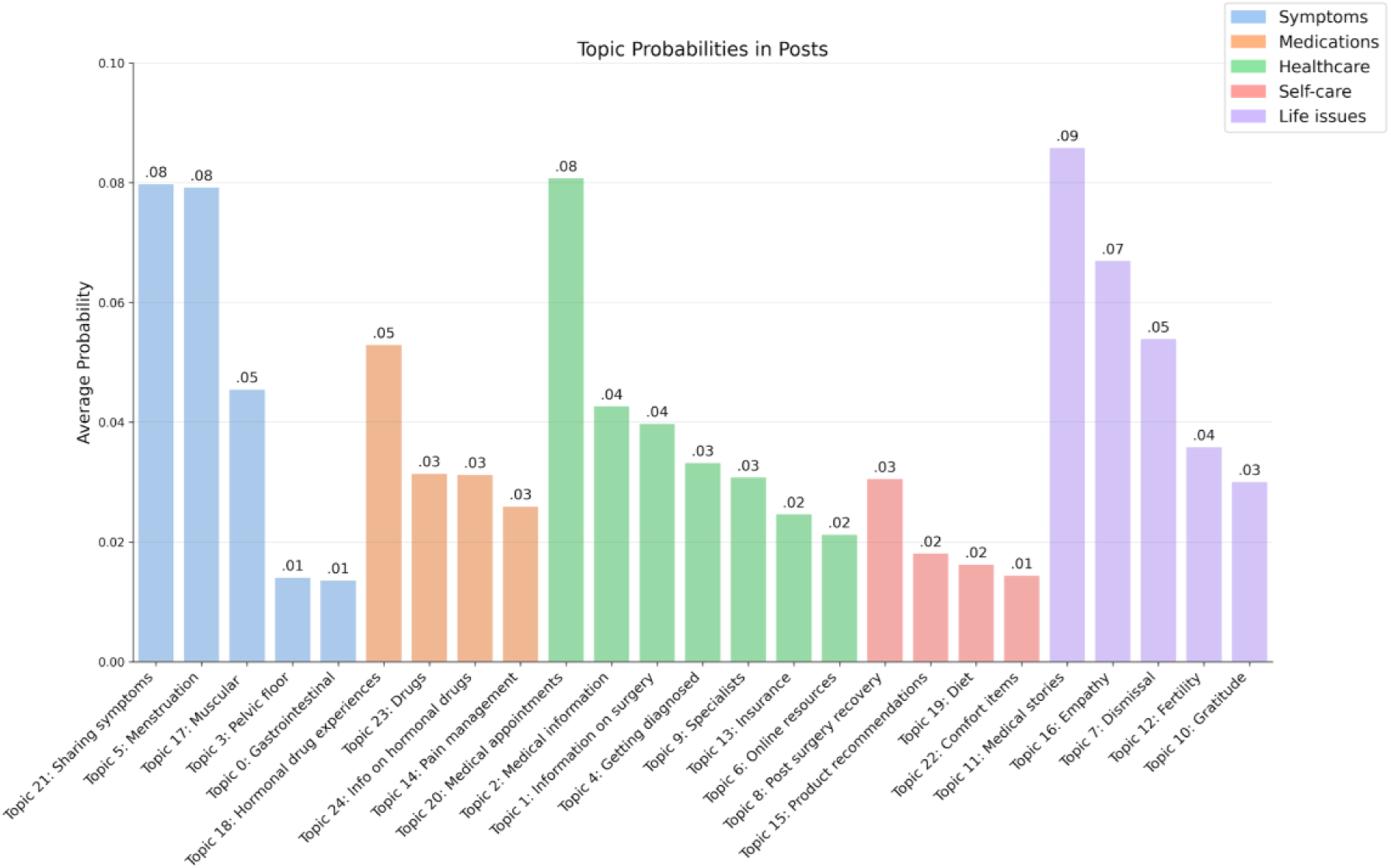
Average topic probabilities in posts collected from the two endometriosis OHCs ordered by the categories. *Medical appointments*, *medical stories*, *sharing symptoms*, *menstruation*, and *empathy* have the highest average probability.

We find that *medical stories* and *medical appointments* are the two most discussed topics. New or returning users frequently recount their healthcare journey at the beginning of their posts: from having the first symptoms as teens, to undergoing surgery, and choosing between treatment options. Other times, users ask specific questions on how to book their medical appointment, what to do if an appointment is moved or the physician does not show up, and what strategies others use to communicate successfully with their doctors.

Two *symptoms* topics, *sharing symptoms* and *menstruation*, are among the most present topics. Users of the endometriosis OHCs share detailed accounts of all their symptoms in order to gain their peers’ opinions on whether they should seek urgent care, whether a new symptom might be caused by their treatment rather than endometriosis, and whether what they are going through resembles other people’s endometriosis.

A large number of posts in the OHCs are solely dedicated to describing menstrual symptoms. New users of these communities are often undiagnosed teenagers who wonder whether they should seek medical assistance given their experiences with menstruation. Furthermore, endometriosis is typically treated with hormonal medicines, which cause additional changes to patients’ menstrual cycles. Patients share such changes with peers to understand if the treatment has been effective at relieving their pain.

*Empathy* is the fifth most present topic in posts of the two OHCs, underlining that demonstrations of empathy are extremely valued by endometriosis patients. Sadly, users often lament feeling misunderstood and dismissed.

### RQ2: What aggregate needs emerge from the OHCs?

In this section, we consider the needs expressed by members of the OHCs. For each of the topics outlined in RQ1, we consider 1) which topics are more likely when different personas are mentioned and 2) what the intent of posts are when they mention each topic. By doing so, we can better understand the interplay between endometriosis experiences, interpersonal relationships, and the goals of OHC members.

#### Personas in the OHCs

Of the four persona categories, posts to the endometriosis OHC most often mention the *endometriosis OHCs*, followed by *medical professional*, *family*, and *partners* (Figure 4).

**Figure 4.**
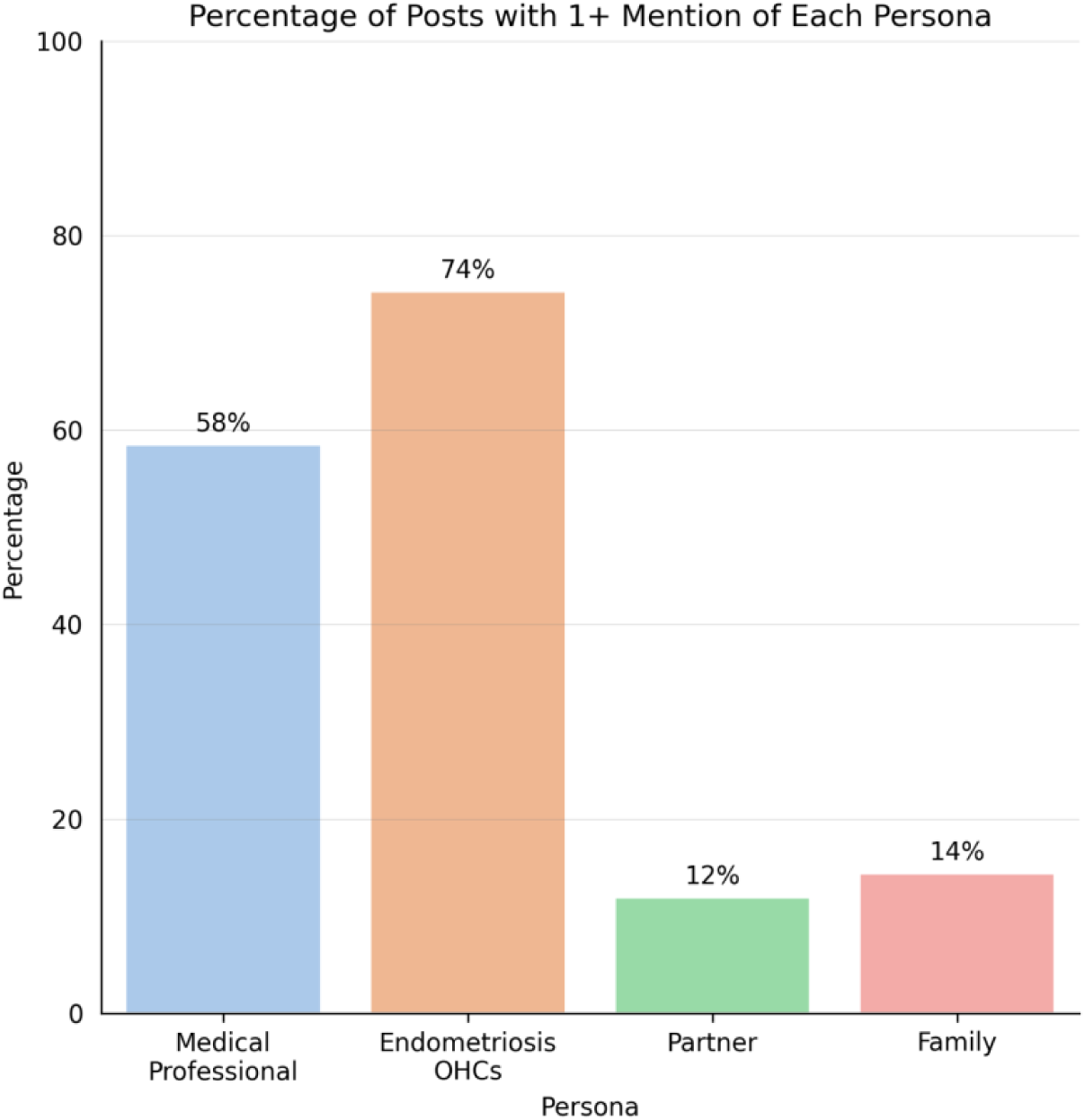
Percentage of posts with more than one mention of each persona in the endometriosis OHCs.

Of posts predicted with at least one of the four personas, we find which topics are most present. For each persona, we find the average topic probabilities for all posts predicted with each persona, converted to *z*-scores. Figure 5 displays this result, depicting what members of the OHCs are most likely to discuss when they mention each persona.

**Figure 5.**
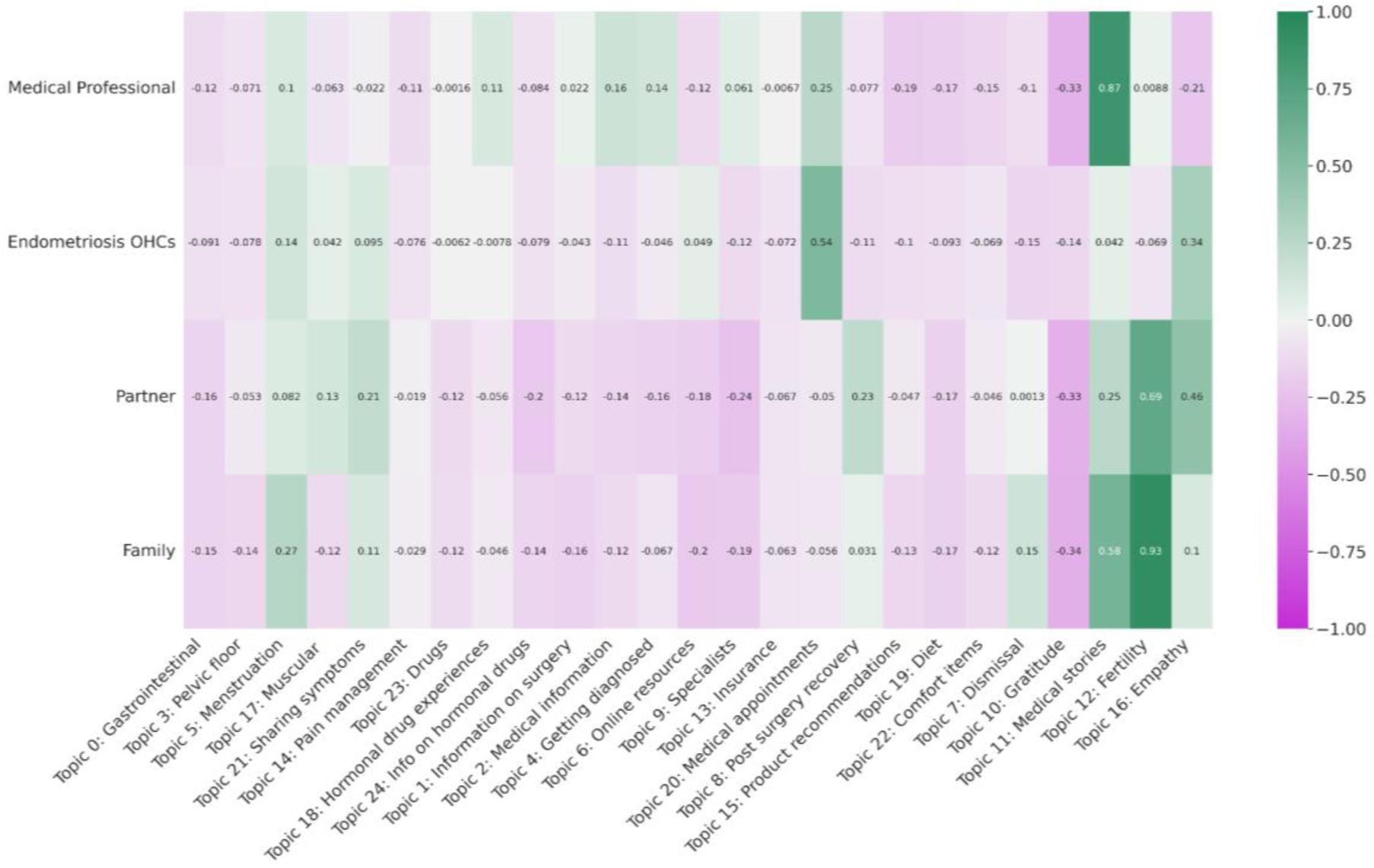
Average topic probabilities (converted to *z*-scores) for posts with different *personas*.

When a *medical professional* is mentioned, posts are more likely about *medical appointments* and *medical stories*, highlighting the important role that providers have in shaping patient medical pathways. However, *medical professional* is the least likely of any persona to be discussed in combination with *empathy* (*P*<.001).

Interestingly, posts with the *endometriosis OHCs* are more likely to discuss *medical appointments* than posts with *medical professional* (*P*<.001). In alignment with our findings in RQ1, users of the OHC request the assistance of the community to prepare for visits, as this support might not be available to them in clinical settings.

Posts that mention *partner* or *family* are likely to discuss topics from the *life issues* category, in particular *fertility* (*P*<.001). These posts emphasize how navigating fertility deeply affects relationships. Mentions of *family* in posts about *fertility* may have to do with family planning and personal goals in growing a family. Some may express concern about being able to have or keep a partner when dealing with infertility. These posts also mention feeling pressure to have children from family or partners.

Lastly, posts that mention *partner* often also discuss *post surgery recovery* (*P*<.001). Partners can indeed play an important role in helping endometriosis patients access treatment and maintaining self-care routines. In addition, it is sometimes the partner of a person with endometriosis who asks for advice from the OHC.

### Intents in the OHCs

We then consider the goals of members of the community in their posts, through our intent predictions. Across all posts, we find that users are most likely to *seek experiences* from the OHC; they do so in roughly half of posts. *Seeking informational support* occurs in around a quarter of posts, and *seeking emotional support* and *venting* are the least common intent types, based on our model (Figure 6).

**Figure 6.**
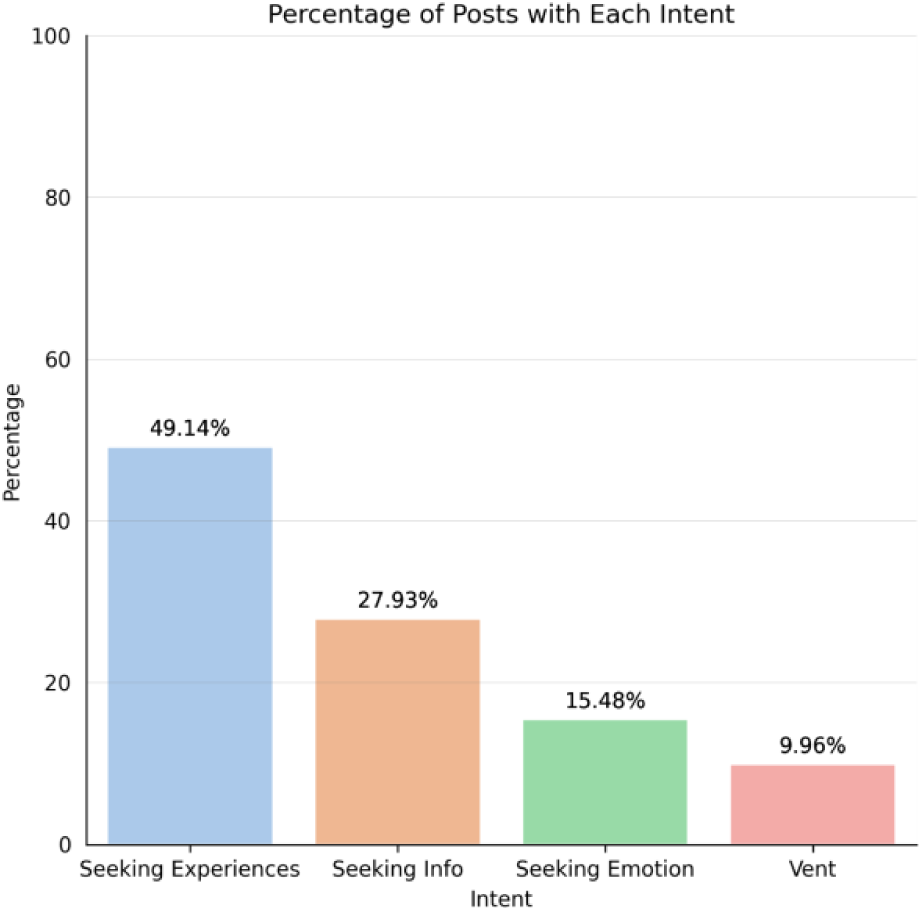
Percentage of posts with each intent label in the endometriosis OHCs.

We then find the average topic probabilities for posts with each predicted intent category. By doing so, we can find which subjects are most related to different goals. When a member seeks information from the community, what are they trying to learn about? When a member simply wants to vent, what subjects are most often related to their frustration?

We find an important divide between the subject matter of posts that *seek experiences* or *informational support* and those that *seek emotional support* or *vent* (Figure 7). The subject matter of posts that *seek information* or *experiences* are more often about topics in the *symptoms*, *medications*, and *healthcare* categories. While members more likely *seek emotional support* and *vent* about the *life issues* topics, including *dismissal*, *medical stories*, *fertility*, and *empathy*.

**Figure 7.**
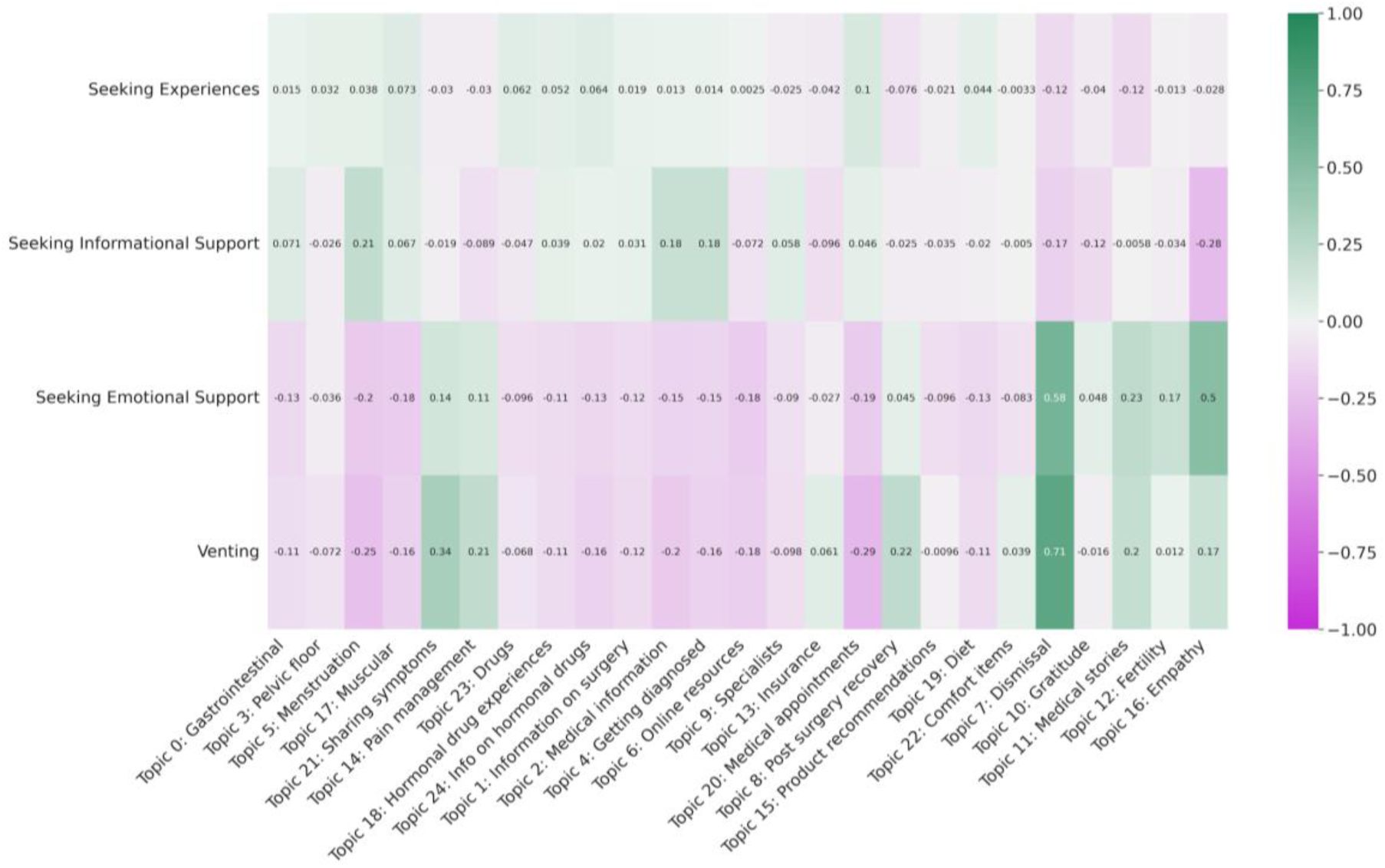
Average topic probabilities (converted to *z*-scores) for posts with different *intents*.

However, members of the endometriosis OHCs do *seek emotional support* – and *vent* – about *pain management* and when *sharing symptoms*. While a person with endometriosis might look for information or experiences regarding their symptoms and pain, they are more likely to look for emotional support from the community or to vent their frustrations.

## Discussion

### RQ1: What aspects of the endometriosis experience are discussed in OHCs

Using topic modeling we find that OHCs are spaces dedicated to narrations of users’ healthcare pathways, directions on how to find care and manage symptoms, as well as expressions of validation between peers regarding their health concerns.

In particular, the most discussed topics in the two communities are *medical stories*, *medical appointments*, *sharing symptoms*, *menstruation*, and *empathy*. These results align with previous findings from qualitative studies collecting endometriosis patients’ experiences. These include evidence of the benefits of sharing one’s story within community [34,59], the need for assistance with treatment regimens and appointments [10,63], the uncertainty experienced by patients related to their symptomatology [11], as well as the value of receiving validation regarding health concerns and symptoms [26].

An existing study of a PCOS subreddit has also found concordance between the OHC user population and research-identified patient cohorts [51]. Although the PCOS OHC includes patients that are typically excluded from clinical trials (such as those with multiple conditions), trends found in laboratory test results posted to the community are consistent with clinically reported results.

Our results also align with studies on OHCs, showing that OHC users become better at communicating with their providers and at managing their conditions, as well as feel less isolated [28,34,35,40,42–44].

### RQ2: What aggregate needs emerge from the OHCs?

Using supervised classification of personas and intents we find that posts mention the endometriosis OHCs more than they mention medical professionals – highlighting the vital role that these groups play in the users’ healthcare decisions –, and that the majority of posts are written to seek experiential advice. Venting is the least common of our intent categories, but venting still occurs in a substantial fraction (10%) of posts.

Combining these classification models with unsupervised topic models, we find that users need assistance with accessing and preparing for medical visits, as well as navigating fertility options. To meet these needs, patients currently turn to the OHCs, their partners, and their family. Interestingly, members of the OHCs seldomly associate medical professionals and providers with empathy.

We also find that patients’ relationships with their partners and family members can be affected by the condition. Users share how physical manifestations of endometriosis, such as infertility, alter their life goals and complicate personal relationships. At the same time, partners and family members play a vital role of serving as informal caregivers. These personas even use the OHCs for advice in creating a strong support system.

Furthermore, while users seek experiential knowledge regarding treatments and healthcare processes, they also wish to vent and establish an emotional connection about the life-altering aspects of the condition.

These results align with previous research on the areas of endometriosis care that need improvement, including non-holistic treatments [1,15,16], unsatisfactory patient-provider communication [5,9,18], and lack of training or educational resources for of patients’ loved ones [15,25–27].

## Conclusions

In this study, we conduct a large-scale analysis of user needs in two endometriosis OHCs, *r/Endo* and *r/endometriosis*. We find that these communities provide members a space where they can discuss care pathways, learn to manage symptoms, and receive validation. Our results also point to the need for greater empathy within clinical settings, easier access to appointments, more information on healthcare processes, and further support to patient loved ones.

Our study demonstrates the value of quantitative analyses of OHCs. OHCs provide very large datasets on patient experiences. In this work, we analyzed hundreds of thousands of posts and comments by tens of thousands of users. This sample size is an order of magnitude larger than that examined in any other study of endometriosis patient needs and experiences of which we are aware. Our results thus fortify findings from small-scale studies about patient experiences and provide insight into hard-to-reach groups.

Lastly, we believe that studies of OHCs can help design interventions to improve care, as argued in previous studies [30,49,51,52].

## Data Availability

All data produced in the present study are available upon reasonable request to the authors.

https://github.com/federicabologna/endometriosis

## Author’s Contributions

FB conducted topic modeling, data analysis, curated visualizations, wrote and revised the manuscript.

RT conducted supervised classification, data analysis, curated visualizations, wrote and revised the manuscript.

KP provided clinical insight into endometriosis symptoms, diagnosis and treatment and revised the manuscript.

MW provided advice and guidance through each step of the project from designing the experiments to revising the manuscript.

## Abbreviations

OHC: Online health community

