## Appendices for "Endometriosis Online Communities: A Quantitative Analysis"

### Appendix A - Fightin' Words Results

We implement Fightin' Words [1] to examine similarities and differences between the vocabularies of r/Endo and r/endometriosis. As represented in Table 1, the two communities have a number of words that distinguish them from each other, suggesting that there may be some differences in norms. However, the words that vary most between the communities (e.g. endometriosis or lap) do not relate directly to our project's research questions and hypotheses, though it is surprising to see some words, including symptoms, distinguish the communities.

Table 1. Fightin' words scores comparing r/Endo (left) to r/endometriosis (right).

| **r/Endo** | | **r/endometriosis** | |
|---|---|---|---|
| **Word** | **z-score** | **Word** | **z-score** |
| lap | 49.64 | short | -30.58 |
| she | 44.56 | pick | -27.48 |
| still | 39.16 | ways | -26.26 |
| didnt | 39.1 | via | -25.67 |
| better | 38.46 | themselves | -25.61 |
| see | 36.29 | system | -25.52 |
| work | 35.7 | wife | -25.43 |
| excision | 35.7 | become | -25.34 |
| find | 35.42 | wanna | -25.28 |
| we | 35.31 | welcome | -24.64 |
| since | 34.66 | head | -24.37 |
| symptoms | 34.49 | entire | -24.04 |
| might | 34.48 | diagnostic | -23.94 |
| people | 34.41 | diarrhea | -23.8 |
| sure | 34.34 | caffeine | -23.72 |
| right | 34.21 | thankfully | -23.66 |
| need | 33.7 | date | -23.49 |
| lot | 33.53 | whether | -23.37 |
| got | 33.28 | besides | -23.33 |
| youre | 33.09 | seemed | -23.19 |

| | | | |
|---|---|---|---|
| make | 32.69 | round | -23.19 |
| things | 32.5 | beginning | -23.17 |
| said | 32.19 | gotten | -22.67 |
| specialist | 31.87 | therapy | -22.39 |
| cant | 31.55 | laps | -22.23 |

In addition, we perform FightinWords on each subreddit against r/PCOS to control for possible errors. We choose r/PCOS since it's an OHC dedicated to discussing a similar severe chronic female genital condition, polycystic ovary syndrome. Interestingly, the similarity between r/Endo and r/endometriosis vocabularies is further supported in this step. As shown in the table below, there is great overlap in the top terms of r/Endo and the top terms of r/endometriosis when compared to r/PCOS. Results are displayed in Tables 2 and 3. In consideration of our hypotheses and the scope of this project, we find that the two subreddits share consistent enough similarities to allow for merging them into a single dataset.

Table 2. Fightin' words scores comparing r/Endo (left) to r/pcos (right).

| **r/Endo** | | **r/pcos** | |
|---|---|---|---|
| **Word** | **z-score** | **Word** | **z-score** |
| pain | 162.26 | pcos | -100.47 |
| endo | 116.64 | weight | -79.57 |
| surgery | 90.12 | hair | -72.01 |
| endometriosis | 66.79 | diet | -48.89 |
| pelvic | 49.07 | acne | -48.36 |
| he | 45.65 | loss | -44.31 |
| specialist | 44.77 | lose | -41.51 |
| lap | 42.17 | low | -38.87 |
| they | 41.23 | eating | -38.67 |
| bowel | 39.28 | high | -38.5 |
| uterus | 38.81 | level | -36.87 |
| cramp | 38.54 | test | -36.41 |
| painful | 37.38 | eat | -36.22 |

| | | | |
|---|---|---|---|
| during | 37.34 | taking | -34.76 |
| laparoscopy | 35.82 | sugar | -34.52 |

Table 3. Fightin' words scores comparing r/endometriosis (left) to r/pcos (right).

| **r/endometriosis** | | **r/pcos** | |
|---|---|---|---|
| **Word** | **z-score** | **Word** | **z-score** |
| pain | 156.76 | pcos | -75 |
| endo | 105.61 | weight | -58.85 |
| surgery | 86.37 | hair | -49.72 |
| endometriosis | 79.02 | diet | -36.18 |
| pelvic | 44.59 | acne | -35.22 |
| cramp | 40.6 | loss | -32.63 |
| painful | 39.11 | lose | -30.42 |
| sex | 38.44 | low | -28.93 |
| they | 38.26 | level | -28.68 |
| bowel | 38.19 | high | -28.64 |
| lap | 37.83 | eating | -28.52 |
| laparoscopy | 37.04 | eat | -27.88 |
| during | 36.36 | taking | -26.9 |
| he | 35.4 | sugar | -25.51 |
| uterus | 34.9 | skin | -25.37 |

### Appendix B - Persona Codebook

B.1 General instructions:

- Label each paragraph based on whether or not the type of persona is mentioned.
- These can be discussions of actual experiences with the given persona, or hypothetical discussions about interacting with a person.

B.2 Medical Professionals

- Includes people who are employed by a medical institution in a patient-facing role.

- Common types of medical professionals include gynecologists, endometriosis specialists, nurses, therapists, etc.

B.3 Family
- Includes anyone in a familial role in position to the person with endometriosis.
- We do not restrict family to immediate family members.
- Partners may be included in this label if they are described as long-term partners. Choose the best option based on context.
- Common types of family members include mothers, fathers, siblings, significant others, children, cousins, aunts, uncles, grandparents, etc.
- At times, people will discuss wanting to have children. Since we include hypothetical discussions of personas, we would include hypothetical children within the family persona.

B.4 Partners
- Includes people who are in a romantic relationship with the person with endometriosis.
- Common types of partners include husbands, wives, SOs, girlfriends, boyfriends, partners, etc.

B.5 Endometriosis Online health Communities
- Includes mentions of the community itself.
- If the post is directed toward the community as an audience and mentions the community while posing questions (e.g. "have you all..."), we include this as a mention of the endometriosis OHC.
- Common mentions of the endometriosis OHC use the terms reddit, subreddit, r/Endo or r/endometriosis, endo warriors, or general phrases like "this community" or "on here"
- Does not include general mentions of a community of people with endometriosis; must specifically be people in the online community.
- For this label especially, consider post context when determining if the endometriosis OHC is or is not present.

### Appendix C - Intent Codebook

C.1 General instructions:
- Many posts explicitly restate their intent at the bottom of the post (and sometimes in the introduction) – whenever possible, use the explicitly stated intent

- Many posts have to share medical information or histories as a way of getting to their intent. In this case, the intent isn't necessarily about sharing medical information
- Posts should typically have one to two intents
    - Some examples:
        - A post may ask multiple questions: if one is informational and the other is for experiences, that would be 2 intents
        - A post may mostly vent but also periodically ask for information, that would also be two intents
- The hardest part will probably be separating seeking information from experiences & separating seeking emotional support from venting

C.2 Seeking Informational Support

- Asking for fact-based information that could just as easily be asked to a medical professional
    - Helpful heuristic: could someone ask their doctor this question?
- Asking about medical details, whether a set of symptoms sounds like endometriosis, etc.

C.3 Seeking Experiences

- In ways, this is the inverse to "seeking informational support"→these are the questions you can't ask doctors
- Asking for descriptions about experiences with surgery, diagnosis, treatment, medicine
- Asking for advice based on personal experiences (what has/hasn't worked for others)
- Asking whether other people have experienced something ("has anyone had X symptom")

C.4 Seeking Emotional Support

- Explicitly looking for comfort or peace of mind
- Sharing positive news and asking for people to celebrate with them
- Posting to the community because no one else in their life "understands"
- Asking for advice about dealing with emotional problems in their relationships, how to not be anxious about their condition/surgery, etc.
- This label has a wider range of positive to negative emotions
- *Note: for this label, look for explicit requests for comfort/peace of mind. Don't project "seeking emotional support" just because someone is frustrated–they might be frustrated and just want medical information*
